# Metabolomic fingerprinting for biomarker discovery in renal amyloidosis

**DOI:** 10.1101/2022.09.21.22280214

**Authors:** Shreya Ghosh, Praveen Singh, Samir Govil, Chayanika Kala, Shivani Chitkara, Shantanu Sengupta, Ashwani Kumar Thakur

## Abstract

Nephrotic syndrome (NS) manifested by proteinuria is the primary clinical hallmark of amyloid deposition in the kidney. However, proteinuria is observed in other kidney disorders misleading clinicians and limiting the scope of early diagnosis. We presumed that amyloid-driven pathophysiology could result in the perturbation of downstream cellular and metabolic pathways in these patients with NS. Herein, we have diagnosed one hundred patients with clinical evidence of nephrotic syndrome. Further, the histopathological evaluation identified the presence of amyloid in eleven patients. To decipher the downstream effects underlying amyloid formation, we performed plasma metabolomic profiling of these patients. Fifteen metabolites, including different lipids, carnitines and amino acids, were found altered in the renal amyloidosis patients compared to controls. It is the first study depicting the potential of metabolomics to identify biomarkers for early diagnosis of renal amyloidosis patients.

## Introduction

Renal amyloidosis involves the deposition of insoluble protein-derived amyloid fibrils in the interstitial spaces, renal cortex and glomerulus of the affected kidney^1^. As the disease progresses, the amyloid fibrils get deposited along the walls of the peripheral glomerulus capillary, resulting in the loss of the filtration barrier for proteins. Consequently, proteinuria ranging from sub-nephrotic to nephrotic is evident in most renal amyloidosis patients depending on the amyloid deposition site^1, 2^. Renal amyloidosis accounts for the majority of cases with nephrotic syndrome in Asian countries, especially among Indian cohorts^3-7^.

The diagnosis is often challenging for clinicians due to its misleading symptoms overlapping with other kidney-associated complications^8^. The histopathological signatures on the affected tissues are the only clinical hallmarks considered for confirming diagnosis^9, 10^. However, associated complications and lack of expertise limit the scope of biopsy-based diagnosis in most cases^11, 12^. We anticipated that amyloid-associated pathologies underlying nephrotic syndrome could perturb the downstream cellular pathways. Based on this, we aimed to track these disordered physiological processes that might play an essential role in discovering a biomarker for early diagnosis. Cellular pathways are the primary outcome of several interactions between biomolecules, including proteins and metabolites. Different multi-omics approaches like genomics, transcriptomics, proteomics and metabolomics are utilized to capture these interactions^13^.

Metabolomics has emerged as a robust tool for identifying the functional relevance of small metabolites, critical components of vital cellular pathways^14^. The untargeted metabolomic approach is preferred over targeted, owing to the unbiased identification of known and unknown metabolites with a broader coverage^15, 16^. In recent years, mass spectrometry has evolved as a promising analytical platform for evaluating the differential expression of these metabolites. It primarily involves the extraction of metabolites followed by their separation, acquisition of mass signatures for individual metabolites and pathway validation^17^. In this study, we have utilized the high-throughput mass spectrometry-based untargeted approach to decipher the downstream effects underlying amyloid formation.

## Materials and methods

### Sample collection

In a two-year single-center study from 2018 to 2020, two hundred patients with renal impairment were screened. Out of these, a hundred patients presented with clinical evidence of nephrotic syndrome. For further evaluation, the renal biopsy specimen was obtained from these patients from the cortex region of the affected kidney (after taking the informed consent). In addition, 2ml of blood was drawn from these patients and controls (having normal renal function) respectively in an EDTA-containing vacutainer. Plasma was collected from these blood samples by centrifugation (1000 x g for 10 minutes at 4ºC). The ethics approval for this study was obtained from the human ethics committee of IIT Kanpur and GSVM medical college (**Ref No** IITK/IEC/2017-18 II-4 & EC/BMHR/2020/18).

### Histopathological analysis of patient

The renal tissues from each suspected patient were obtained via ultrasound-guided renal biopsy. The fresh tissues (collected in saline solution) were then processed, followed by paraffin embedding. Finally, a series of staining procedures were performed to identify the pathological hallmarks.

#### Hematoxylin and Eosin staining of the FFPE tissue sections

5μm thick deparaffinized tissue sections were rehydrated serially in different grades of ethanol (100% to 70%), followed by final rehydration in distilled water. The hydrated sections were then put in Hematoxylin-Mayer’s solution (Sigma Aldrich, catalog number: H9627) for two minutes, followed by a brief wash in distilled water and counterstaining with 1% eosin Y (Merck Millipore, catalog number: 115935) solution for thirty seconds. It was then subjected to subsequent dehydration and xylene treatment. Finally, the slides were mounted in mounting media and observed in bright light under a microscope.

#### Periodic acid Schiff staining of the FFPE tissue sections

The patient-derived renal tissue sections were deparaffinized and hydrated with deionized water. It was then immersed in 0.5% periodic acid (Himedia, catalog number: GRM1837) solution for five minutes at room temperature (18-26ºC), followed by rinsing in different changes of distilled water. Next, the sections were immersed in Schiff’s reagent (Merck Millipore, catalog number: 109033) for fifteen minutes at room temperature, followed by washing them in lukewarm warm water for five minutes. Finally, it was counterstained in hematoxylin solution for ninety seconds and then rinsed in running tap water for 5 minutes. The slides containing stained tissue sections were then dehydrated and mounted in mounting media, followed by visualization under the microscope in bright light.

#### Congo red staining of the FFPE tissue sections

The stock and working solutions of Congo red (Sigma Aldrich, catalog number: C6277) were prepared according to the published protocols ^18^. Deparaffinized tissue sections were stained with hematoxylin solution for two minutes. It was then rinsed with tap water for two minutes and then in distilled water twice for two minutes each. The sections were then placed in Congo red working solution A for twenty minutes. Next, they were transferred to the Congo red working solution B for twenty minutes. The sections were then rinsed briefly in two changes of absolute ethanol for ten seconds each. Finally, the sections were cleared of leftover ethanol in two changes of xylene followed by mounting the slide under the coverslip in synthetic DPX medium.

### Plasma Metabolomic profiling of patients using a mass spectrometry-based platform

#### Extraction of metabolites from Plasma

400 μl of methanol (Biosolve, catalog number: 136841) was added to 100 μl of plasma, vortexed for 15 seconds, and incubated on ice for 1 hour to allow precipitation of blood proteins. It was then centrifuged in 15,000 x *g* for 15 minutes at 4ºC to remove the precipitated proteins. The supernatant was then lyophilized, followed by reconstitution in 50% methanol for further LC-MS analysis.

#### Separation of extracted metabolites by reverse phase HPLC

The reverse phase chromatography was carried out on an Ultimate 3000 UHPLC system (Thermo Scientific). The two mobile phases used for separation consisted of water LC-MS-grade, 10 mM ammonium acetate with 0.1% formic (*v/v*) acid (Sigma Aldrich, catalog number: 09676) (buffer A) and acetonitrile (Biosolve, catalog number: 012041) with 0.1% formic acid (*v/v*) (buffer B). The extracted metabolites were initially re-suspended in 75 μl of 50% methanol and were separated on a C18 Column (ACQUITY UPLC BEH Amide Column, 130Å, 1.7 μm, 2.1 mm X 100 mm, Waters) with a column oven temperature of 30□. The flow rate was set to 200 μl per minute. The linear mobile phase B gradient was applied from 5% to 95%B. The following gradient program was employed: 5% buffer B (100% acetonitrile with 0.1% formic acid) for 1 min, 5% to 40% buffer B in 2 min, 40% buffer B for another 2 min, 40% to 95% buffer B in 10 min, 95% buffer B for 5 min, 5% buffer B in 6 min and 5% buffer B is kept constant for next 5 min. This resulted in a total gradient time of 30 min.

#### Separation of extracted metabolites by hydrophilic interaction chromatography

The HILIC separation was also performed on the Ultimate 3000 UHPLC system (Thermo Scientific), maintaining column temperature at 30ºC. The mobile phase A was water LC-MS grade with 10 mM ammonium acetate and 0.1% formic (*v/v*) acid. While the mobile phase B was 10 mM ammonium acetate solution in 90% LC-MS-grade acetonitrile with 0.1% formic acid (*v/v*). The flow rate was set at 300 μl per minute. A linear gradient of mobile phase B was applied from 95% to 10%. The gradient was as follows: 95% buffer B (90% ACN, LC-MS-grade with 10 mM ammonium acetate with 0.1%formic) for 0.5 min,90% buffer B in the next 1 min, 50% buffer B in the next 7 min, 30% buffer B in another 0.5 min, 10% buffer B in 1 minute which is held constant for 2.5 min, buffer B was finally brought to 95% in next 1 min which is kept constant for next 1.5 min. This resulted in a total gradient time of 15 min.

#### Acquisition of ESI-MS and ESI-MS/MS data

The acquisition was performed on a QTOF mass spectrometer (TTOF 5600+, SCIEX) equipped with an electrospray ionization source and coupled to an Ultimate 3000 UHPLC system (Thermo Scientific). All the samples were scanned over a mass range of 50–1000 *m/z* for both the reverse phase and HILIC and were analyzed in both positive and negative modes to ensure complete coverage of metabolome ^19^. The mass spectrometer was operated in a data-dependent manner. The *m/z* peaks having a mass intensity greater than 100 and charge +1 were further subjected to MS/MS fragmentation in each duty cycle. The collision energy was set at 30 eV with an energy spread of 15 eV.

#### Processing of raw MS-spectral data

Peak finding and alignment across the samples were performed using MarkerView v1.2.1 (SCIEX). The peak finding criteria were set as follows: minimum spectral peak width was set to 5 ppm, minimum RT peak width was set to 5 scans, noise threshold of 500 was used, and charge state was assigned. For peak alignment, retention time tolerance of 1 min and mass tolerance of 5 ppm was used. Peaks present in less than 70% of samples were filtered out. Only monoisotopic peaks were used for quantification. Data was exported in a spreadsheet format for further analysis.

#### Statistical analysis

The observed metabolic features were subjected to multivariate statistical analysis using Metaboanalyst 5.0 software ^20^. Initially, the spectral intensities were normalized based on the sum of peak area to minimize data variability and make the samples more comparable ^21^. Percentage relative standard deviation (RSD%) was calculated on QC samples to assess variability from extraction. Metabolic features having RSD >40% on the QC samples were excluded before statistical analysis ^22^. Further, the normalized data were log-transformed and auto-scaled. Intrinsic clustering between the samples was evaluated using unsupervised partial component analysis (PCA). Supervised partial least square discriminant analysis (PLS-DA) was done to determine the important metabolic feature that can significantly cluster and differentiate controls from cases ^23^. In addition, the validation and predictive accuracy of the PLS-DA models were estimated using calculated R-square and Q-square values. Univariate analysis like fold change test, t-test and volcano plot was done to determine the panel of differential metabolites between the two groups ^24^. Besides, variable importance in the form of VIP score was calculated based on these PLS-DA models.

#### Identification of differentially altered metabolites

The panel of metabolic features having VIP scores greater than one was identified in MasterView software (SCIEX) using SCIEX-an all-in-one HR-MS/MS library with NIST 2017 library bundle. For metabolic feature not identified with the above library, MS search and structure matching was performed with the metabolite entries in databases like the human metabolome database (HMDB), METLIN and LIPID MAPS in PeakView v2.2 (SCIEX).

## Results

The clinical hallmarks of nephrotic syndrome are the urinary excretion of excess proteins, decreased blood albumin levels, and edema^25^. Similar symptoms and biochemical parameters were reflected in hundred out of two hundred patients screened in the study. Besides, hepatomegaly and anasarca were observed in some of the patients. Histopathological examination of using light microscopy showed diffuse mesangial hypercellularity in the glomerulus (Figure S1 A, Supplementary information). In addition, interstitial fibrosis, characterized by the presence of eosinophilic hyaline-like deposits, was evident in the interstitial spaces (Figure S1 B, Supplementary information) of these patients. Further, distended tubules with mild thickening in the tubular basement membrane were also seen (Figure S1 C).

### Amyloid deposition in the kidney of patients with nephrotic syndrome

Amyloid deposits are well known to appear as amorphous hyaline-like material in the affected tissues ^26^. Deposition of amyloid in the tubulointerstitium causes interstitial fibrosis and tubular atrophy, characterized by the thickened tubular basement membrane^2^. In addition, renal amyloid deposits are accompanied by mesangial hypercellularity in some cases ^27, 28^. These histological signatures in the renal biopsy sections of fifteen patients heightened our suspicion of amyloid presence. Thus, classical Congo red (CR) staining was performed on these renal tissue specimens to validate the observation. Hyaline-rich deposits in the glomerulus and interstitial spaces (Figure S2 A, Supplementary information) of eleven renal biopsies exhibited apple-green birefringence under polarised light (Figure S2 B, E, Supplementary information). Upon changing the angle of the polariser by 10^º^ either in a clockwise or anticlockwise direction, a transformation in the birefringence from apple green to either bluish-green or reddish-orange (depending on the orientation of fibrils) was evident (Figure S2 C, D, F, Supplementary information). Clinicians and pathologists use this property of CR to detect amyloid deposits in pathogenic tissue specimens and reduce ambiguity.^29, 30^. Thus, it confirmed the amyloid nature of eosinophilic deposits in eleven patients.

Progression of amyloid formation in any disease sometimes occurs due to metabolic imbalances^31, 32^. Besides, differential expression of metabolites is reported in systemic amyloidosis affecting the heart and nervous system^22, 33^. However, the perturbed metabolome underlying amyloid formation in the kidneys has not been explored to date. Thus, we aimed to track the metabolic perturbations to get insights into the amyloid-associated pathophysiology. The plasma samples collected from these biopsy-proven amyloid positive eleven patients and fifteen controls (Table 1), were subjected to metabolomic profiling (Figure 1).

**Table 1.** Renal function test profile of the study participants

| Characteristics | Mean values ± SD | | Reference values |
| --- | --- | --- | --- |
|  | *Cases* | *Controls* |  |
| Age (years) | 34 ± 11.05 | 43±14.50 | |
| 24-hour urine protein (gm/24hr) | 4 ± 2.23 | ND | <1gm/24 hr |
| Serum creatinine (mg/dl) | 1.6 ± 0.81 | 0.84±0.20 | 0.5-1.5 |
| Serum albumin (gm/dl) | 2.5 ± 0.94 | 4.22±0.39 | 3.5-5.2 |
| Serum urea (mg/dl) | 74 ± 39.76 | ND | 10-45 |
| Ionic calcium (mg/dl) | 4.58 ± 0.44 | ND | 4.5-5.5 |

**Note:** ND- not determined, gm- gram, hr- hour, mg- milligram, dl- decilitre, SD- standard deviation.

**Figure 1.**
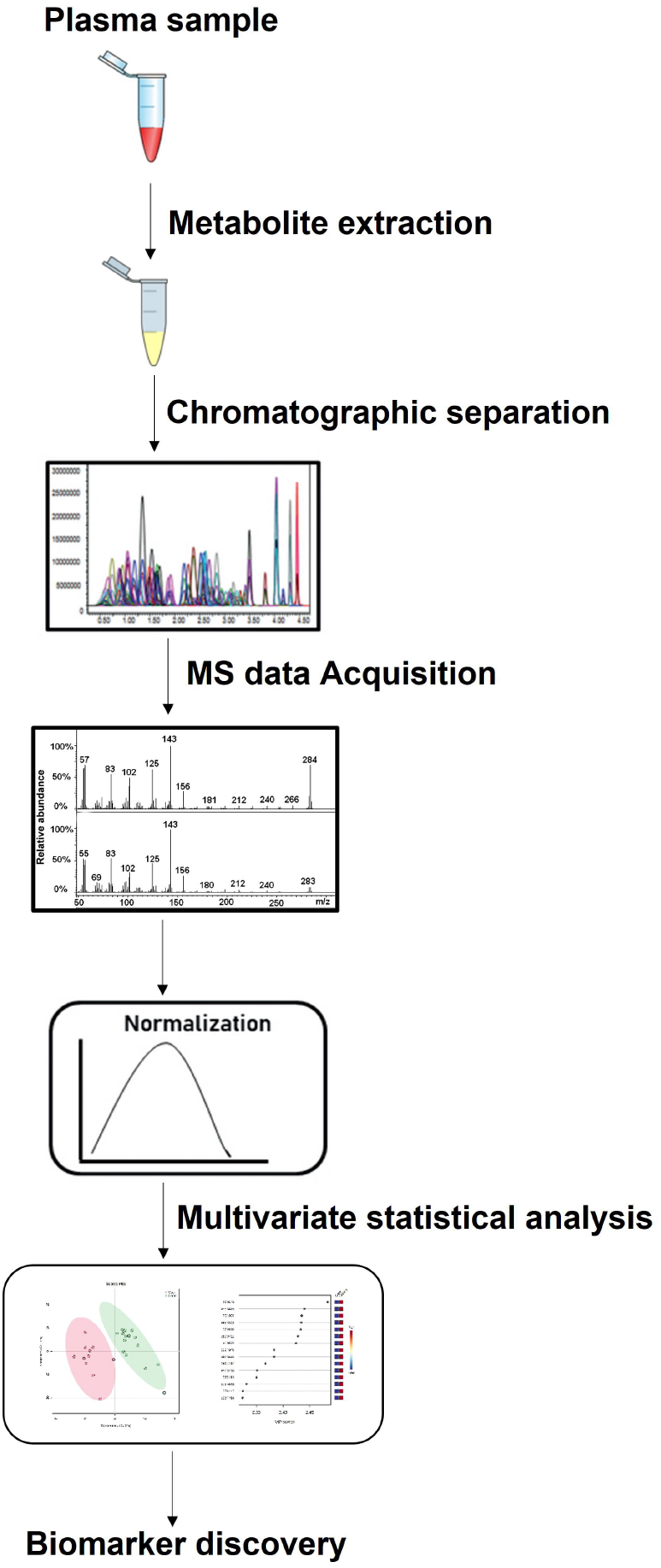
Schematic representation of the workflow for metabolomic fingerprinting.

### Metabolomic analysis

#### Analysis of the mass spectrometry data

One of the important attributes of using an untargeted metabolomic platform is the ability to replicate each LC run for different biological samples. The differential expression is based on the peak intensity for each m/z (metabolic feature) in an untargeted approach. Thus, it is necessary to produce a reproducible chromatogram for such analysis. Herein, the total ion chromatograms extracted from PeakView software (SCIEX) for both the HILIC and reverse phase analysis was reproducible for all the samples (Figure S3 Supplementary information). The spectra of one control sample were found to be abrupt and were excluded for further statistical analysis.

#### Statistical Analysis

Multivariate analysis of the metabolites using unsupervised principal component analysis (PCA) showed a clear demarcation in the observed features between cases and controls (Figure 2A). A maximum variance of 23.8% was observed in the first principal component, and a minimum variance of 6.2% in the fifth component (Figure 2B). In addition, supervised PLS-DA analysis showed differential clustering among the two groups (Figure 3A) with a maximum variance of 18% and a minimum variance of 7.9% (Figure 3B). The model’s predictive accuracy tested with 10-fold cross-validation showed a good fit of the PLS-DA model (Figure 4A) with R square and Q-square greater than 0.7 (Figure 4B). Fold change analysis with t-test identified a total of 17 and 122 metabolic features significantly to be upregulated and downregulated, respectively, in patients as compared to controls (Figure 5A). Based on the calculated variable importance projection (VIP) score (using PLS-DA models), 80 metabolic features (m/z values) were noted to be altered.

**Figure 2.**
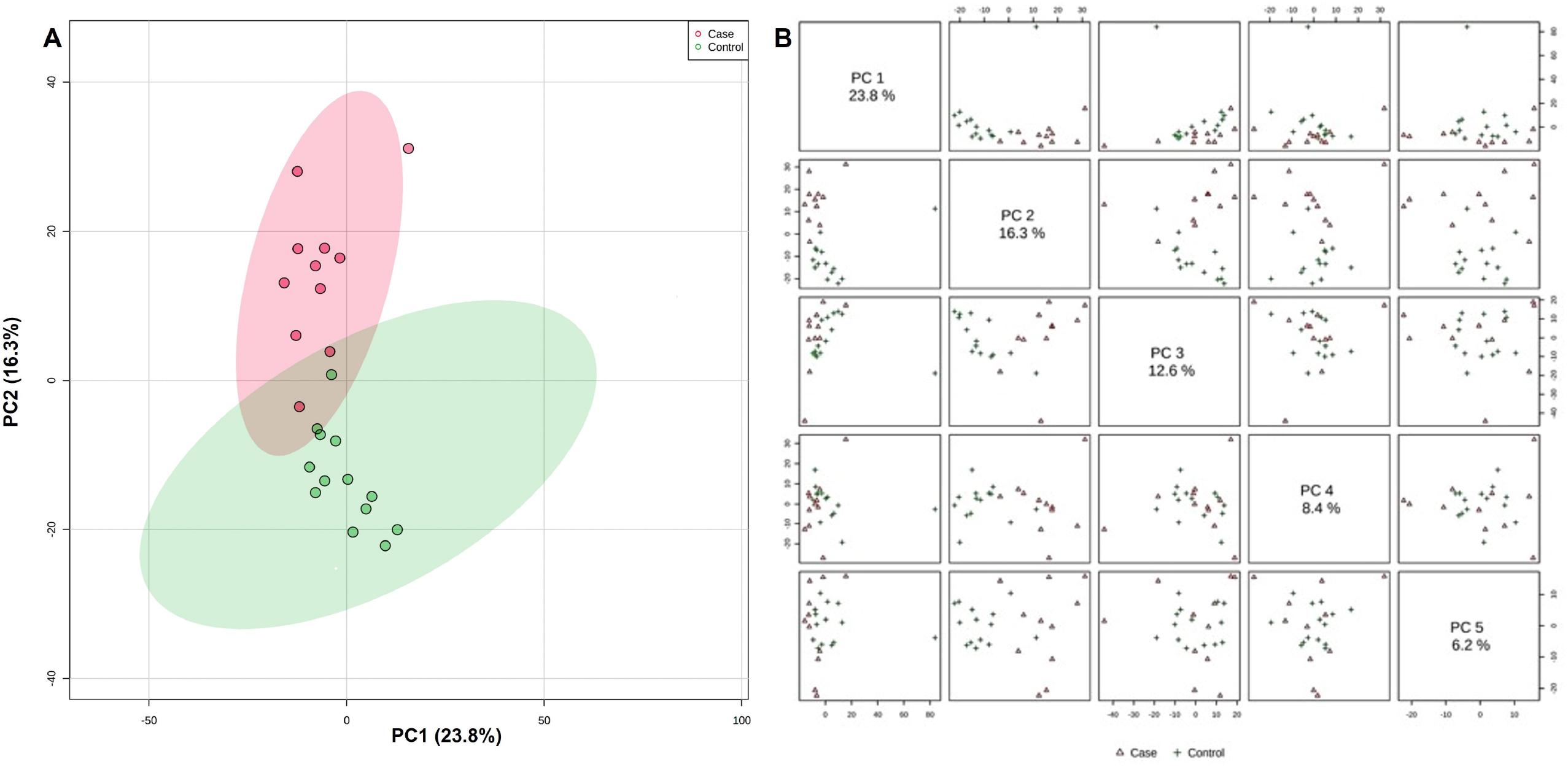
Unsupervised PCA plot depicting differential distribution of metabolic features among two groups (A). A decreasing trend in variance values was observed in all the components of the principal component analysis (B).

**Figure 3.**
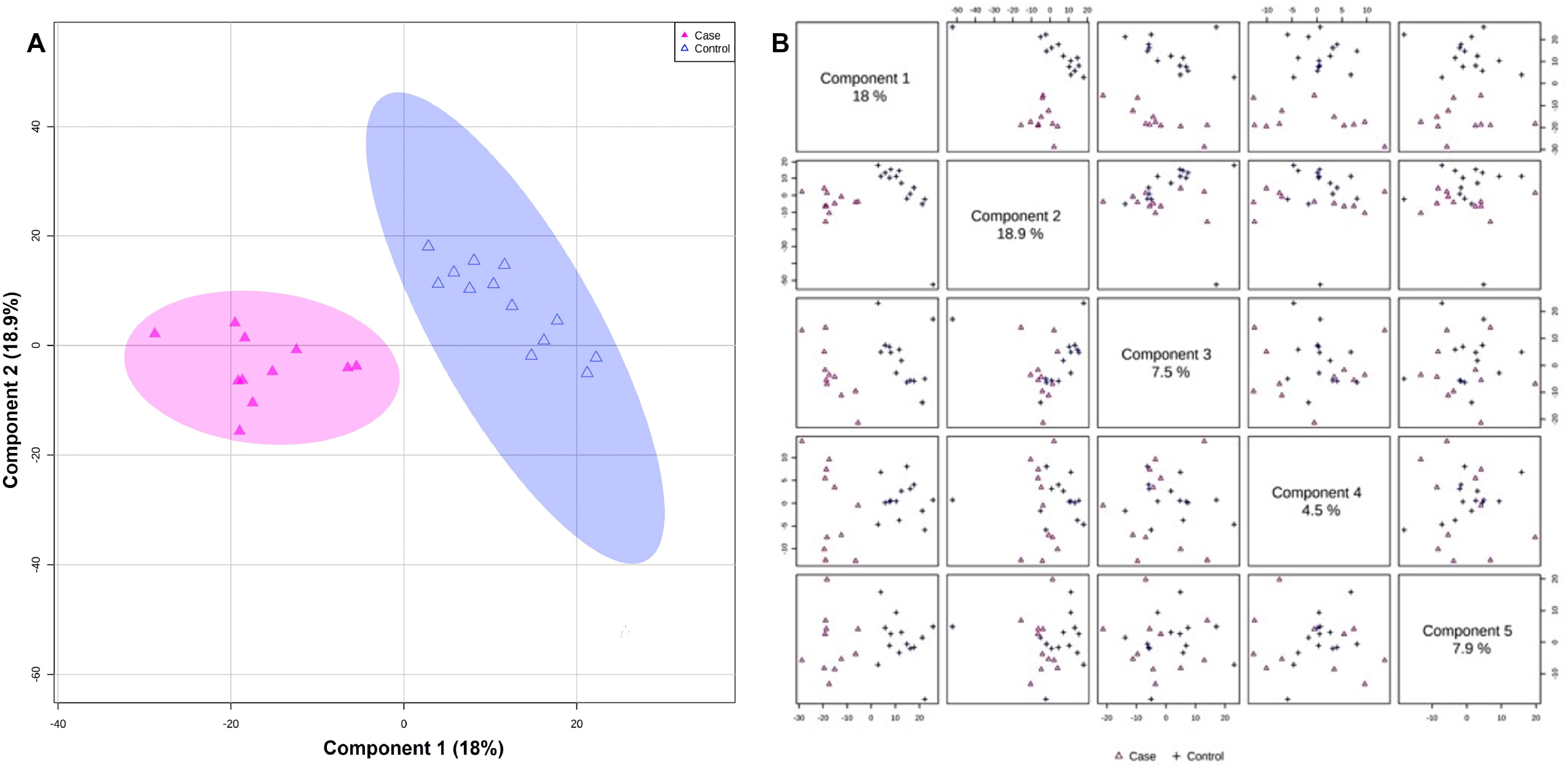
Supervised partial least square discriminant analysis (PLS-DA) model discriminating the metabolic features into two groups (A). A similar decreasing trend in the variance was noted in all the components of the model (B).

**Figure 4.**
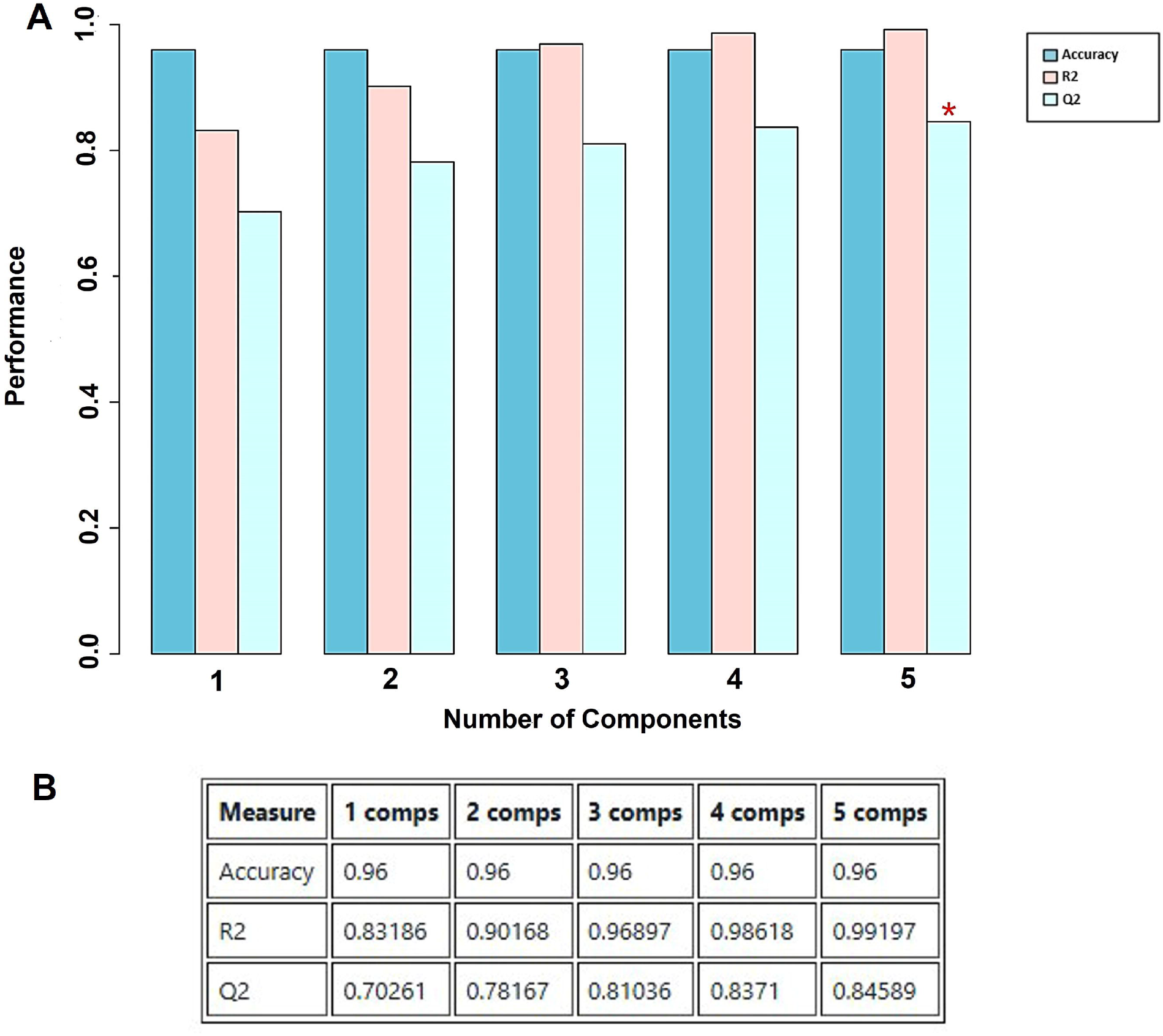
Predictive ability of the PLS-DA model based on 10-fold cross validated Q2 values (A) showing accuracy, R-square and Q-square values of greater than 0.7 (B). This indicated the reliability of the model in predicting the metabolic changes among the two groups. The red star in A represents the best classifier model.

**Figure 5.**
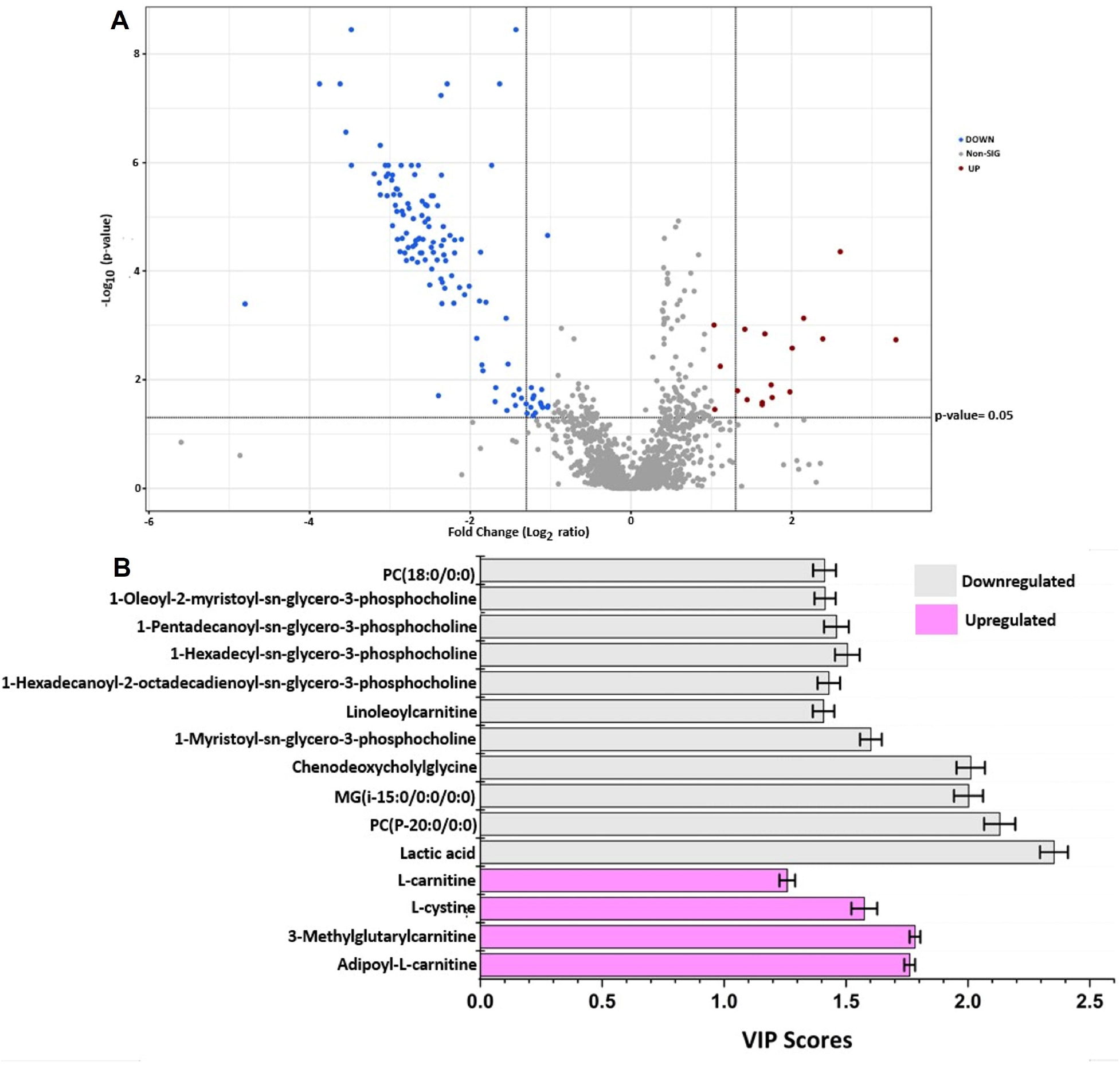
Volcano plot analysis depicted differentially expressed metabolic features (A) in patients as compared to controls (A). The variable importance projection score plot highlighted important 15 metabolites with VIP score greater than 1, discriminating patients from controls (B).

#### Differential distribution and expression of identified metabolites among the two groups

A total of 15 biologically important metabolites with a false discovery rate (FDR) less than 0.05 and a VIP score greater than 1 (Figure 5B) were significantly altered between the patients and controls. Initially, all these metabolites were subjected to clustering analysis based on the VIP scores. The generated heat map highlighted their differential expression and clustering among cases and controls (Figure 6). Besides, these metabolites showed differential distribution among the two study groups (Figure S4, Supplementary information). An increased concentration of L-cystine and different carnitines was observed in renal amyloidosis patients compared to controls (Table 2). Similarly, several classes of lipids were found to be downregulated in patients having renal amyloid deposits (Table 3). The chromatographic representation of mass ion intensities from cases and controls for some of these altered metabolites identified in this study also reflected the same pattern (Figure S5, Supplementary information). This, in turn, validated our observations and analysis.

**Table 2.** List of metabolites upregulated in renal amyloidosis patients as compared to controls

| Metabolite | Observed m/z | Fold Change | p-value | VIP Scores | FDR |
| --- | --- | --- | --- | --- | --- |
| Adipoyl-L-carnitine | 290.149 | 9.8369 | 0.001847 | 1.7611 | 2.81E-04 |
| 3-Methylglutarylcarnitine | 290.149 | 9.8369 | 0.001847 | 1.783 | 2.81E-04 |
| L-Cystine | 239.0141 | 2.1615 | 0.005676 | 1.575 | 8.38E-04 |
| L-Carnitine | 162.1116 | 2.3463 | 0.01324 | 1.25898 | 0.013124 |

**Note:** VIP-variable importance projection score, FDR- False discovery rate, m/z- mass/charge.

**Table 3.** List of metabolites downregulated in the renal amyloidosis patients as compared to controls

| Metabolite | Observed m/z | Fold Change | p-value | VIP Scores | FDR |
| --- | --- | --- | --- | --- | --- |
| Lactic acid | 89.0241 | 0.37065 | 3.58E-09 | 2.3532 | 9.91E-11 |
| PC(P-20:0/0:0) | 536.4099 | 0.13097 | 6.09E-06 | 2.1304 | 7.21E-07 |
| MG(i-15:0/0:0/0:0) | 317.2693 | 0.1609 | 2.46E-05 | 2.0023 | 3.01E-06 |
| Chenodeoxycholylglycine | 467.3509 | 0.21824 | 4.67E-05 | 2.01172 | 5.85E-06 |
| 1-Myristoyl-sn-glycero-3-phosphocholine | 468.3076 | 0.27637 | 0.005357 | 1.6019 | 8.18E-04 |
| Linoleyl carnitine | 424.3245 | 0.46093 | 0.028983 | 1.40784 | 0.004235 |
| 1-Hexadecanoyl-2-octadecadienoyl-sn-glycero-3-phosphocholine | 758.5667 | 0.43265 | 0.01968 | 1.4295 | 0.003068 |
| 1-Hexadecyl-sn-glycero-3-phosphocholine | 482.3246 | 0.46309 | 0.015205 | 1.50536 | 0.002349 |
| 1-Pentadecanoyl-sn-glycero-3-phosphocholine | 482.3246 | 0.46309 | 0.015205 | 1.4607 | 0.002349 |
| 1-Oleoyl-2-myristoyl-sn-glycero-3-phosphocholine | 732.5529 | 0.40485 | 0.027869 | 1.41414 | 0.004162 |
| PC (18:0/0:0) | 568.3622 | 0.38882 | 0.021899 | 1.4119 | 0.003279 |

**Note:** VIP-variable importance projection score, FDR- False discovery rate, m/z-mass/charge.

**Figure 6.**
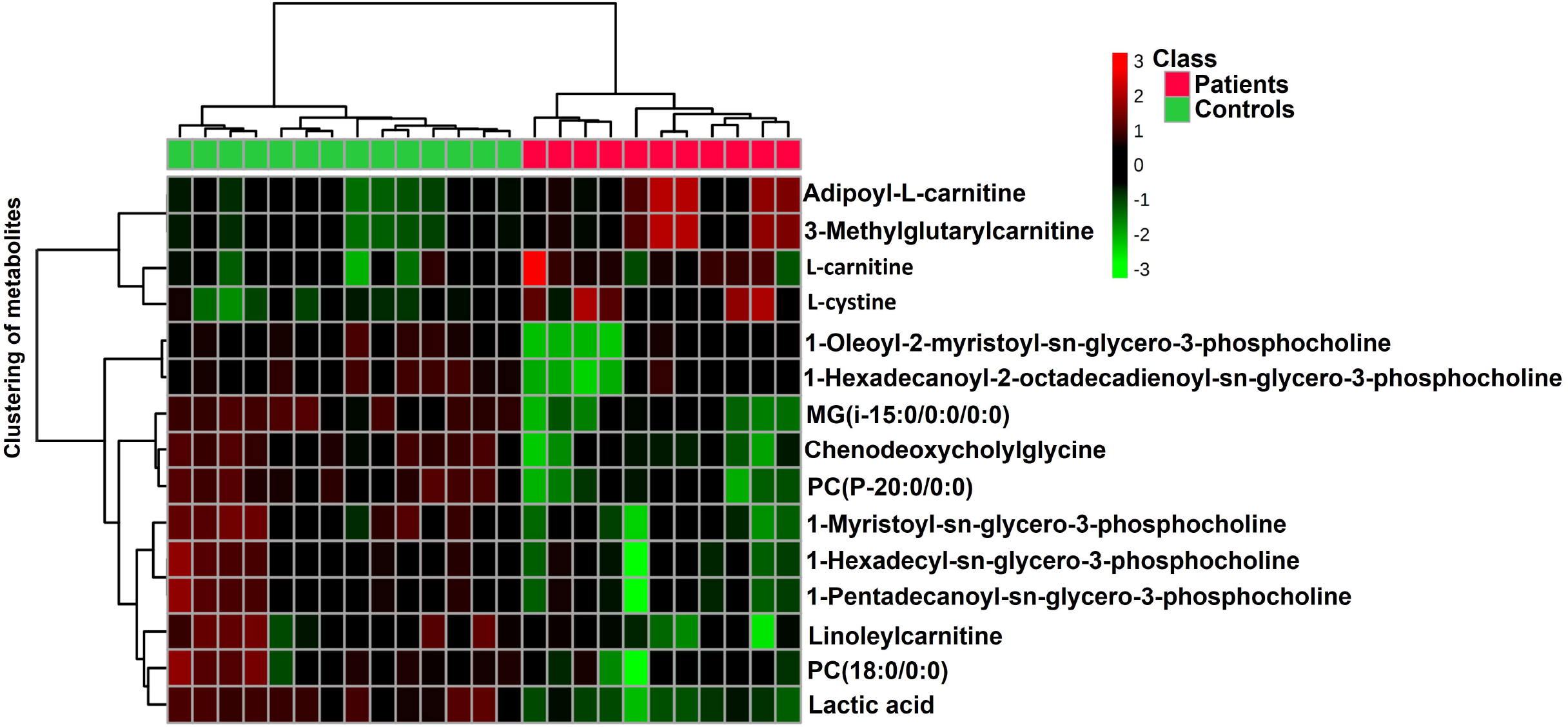
Heat-map analysis showed differential expression of these 15 important metabolites between patients and controls. Each coloured cell on the map represents concentration values of the identified metabolites in different samples.

## Discussion

The present study has depicted amyloid formation as one of the players contributing to the pathophysiology underlying nephrotic syndrome. Amyloid deposition poses damage to the glomerular filtration barrier, resulting in the release of blood and urine proteins^34^. As a result, it causes physiological imbalances and disturbs the downstream biological pathways.

An important diagnostic tool for understanding disease-associated pathologies is the assessment of biological markers ^35^. They are the key indicators of the intricate cascade of molecular events underlying a disease condition. Identifying potential biomarkers in biological fluids promotes a better understanding of disease onset and progression ^36^. Previous studies have showcased the role of biological markers in the early diagnosis of amyloid-associated disorders ^22, 37-41^. Despite the presence of biological markers signifying renal failure and associated diseases ^42, 43^, distinct diagnostic markers for renal amyloidosis have not been explored to date. Thus, we fingerprinted the metabolic signatures in the plama of patients with renal amyloid deposition.

The current study has identified a broad array of fifteen altered metabolites in renal amyloidosis patients presenting with nephrotic syndrome, unlike those reported for NS in the literature^44, 45^. Out of these, L-cystine and different types of carnitine levels were found to be upregulated. In contrast, different lipid classes were found to be significantly downregulated.

Increased levels of acylcarnitines like adipoyl-L-carnitine and 3-methyl glutaryl carnitine were found in patients having renal amyloid deposits. Acyl carnitines serve as the markers for lipid metabolism in mitochondria. Impairment in mitochondrial function causes dysregulation of the fatty acid metabolism, leading to an upregulation in the acylcarnitine levels in blood ^46-48^. Pre-fibrillar amyloid proteins are known to induce mitochondrial dysfunction ^49^. In addition, renal fibrosis often accompanies mitochondrial dysfunction ^50, 51^. In the present work, the presence of amyloid deposits and interstitial fibrosis was evident in the study participants. Thus, we believe that the underlying pathology in these patients might upregulate the carnitine and acylcarnitine levels, suggesting their putative role as future diagnostic markers. However, the elevation of lactate in mitochondrial dysfunction is usually condition-dependent ^52^, accounting for the reduced lactate levels in our cases compared to controls.

Mitochondrial dysfunction predisposes oxidative stress, an imbalance in the formation of reactive oxygen species (ROS) and subsequent antioxidants ^53^. The generated ROS causes oxidative modification of cysteine residues to form L-cystine ^54^, a sensitive marker reported for redox stress ^55^. Oxidative stress is an independent risk factor for amyloid formation and its associated pathogenesis ^56, 57^. Thus, oxidative stress developed in these amyloidosis patients might have caused an increase in the plasma L-cystine levels. Fatigue is reported to be associated with lower levels of linoleyl carnitine in plasma ^58^. Hence, the down-regulation of linoleyl carnitine in this study signifies fatigue condition, an underlying symptom of impairment of mitochondrial function ^59^.

Renal amyloidosis is reported to cause substantial weight loss in the affected patients ^60-62^. One of the critical indicators for weight loss is a decrease in the levels of lysophosphatidylcholine in plasma ^63^. In the present study, reduced levels of different phosphatidylcholines and lysophosphatidylcholines might contribute to the amyloidosis-associated weight loss in these patients.

Monoacylglycerols are the primary breakdown product of triglycerides via lipase enzyme. Chronic renal failure is associated with elevated levels of apoC-III, a known inhibitor for lipase enzyme ^64^. Thus, monoacylglycerol can be a potential candidate for assessing the severity associated with renal impairment in these patients.

In our study, the metabolites of several known drugs and plant-based dietary supplements were also identified, probably due to the presence of non-fasting samples. However, these metabolites were not considered for further statistical analysis to reduce ambiguity. The limitations of this study are: Firstly, the sample size is limited. As it is a pilot study, it needs to be conducted on a large sample size to attain significant clarity. Secondly, all the samples from patients and controls were a mixture of fasting and non-fasting samples, as no record was available from the sample collection center. However, a sub-analysis was performed to trace the dependence of observed metabolome status on the nutritional status of the study participants. Based on this, five of the identified metabolites, mainly di and triglycerides, were excluded after data analysis. It was done to remove the variation from the fasting/no-fasting state of the blood sample ^65^.

## Conclusion

Renal amyloidosis is an underlying cause of idiopathic nephrotic syndrome (INS) in adult patients. However, its early diagnosis is quite challenging for clinicians. In the present study, we have depicted amyloid-associated pathophysiology by capturing the altered plasma metabolome in renal amyloidosis using an LC-MS-based technique. Mass spectrometry-based amyloid typing and proteomic profiling on formalin-fixed tissue samples are the only techniques reported for diagnosing renal amyloidosis patients^66-68^. To the best of our knowledge, this is the first study for biomarker discovery in renal amyloidosis underlying nephrotic syndrome. This work has set the platform for clinicians to diagnose the early onset and progression of amyloid formation in the kidney. Altogether, it has opened new avenues for discovering new biomarkers in the future, relieving patients from the currently used painful biopsy-based diagnosis. Despite having statistical significance, the clinical applications of these metabolites as diagnostic markers need more future studies, including validation in a large number of renal amyloidosis patients.

## Data Availability

All data produced are available online as supplementary information

## Abbreviations

ºC: degree Celsius
μl: microlitre
ACN: acetonitrile
CR: Congo red
EDTA: ethylene diamine tetra acetate
eV: electron volts
FDR: false discovery rate
FFPE: formalin-fixed paraffin-embedded
GD: glomerular disease
GFB: glomerular filtration barrier
HILIC: hydrophilic interaction liquid chromatography
HMDB: human metabolome database
IgA: immunoglobulin A
NS: nephrotic syndrome
INS: idiopathic nephrotic syndrome
LC: liquid chromatography
min: minutes
ml: milliliter
mM: millimolar
MS: mass spectrometry
PAS: periodic acid Schiff
PCA: partial component analysis
PLS-DA: partial least square discriminant analysis
ppm: parts per million
QC: quality control
RFT: renal function test
RSD: relative standard deviation
RT: retention time
TFA: trifluoroacetic acid
TOF: time of flight
US: United States
VIP: variable importance projection

## Acknowledgments

SG is thankful to MHRD and IIT Kanpur for funding her PhD fellowship. PS acknowledges CSIR for funding his PhD fellowship. The authors sincerely thank Dr Ashok Verma for conducting the renal biopsies and Dr. Richa Giri for her scientific inputs during discussions. They also acknowledge the contribution of Dr. Megha Harke Uppin for reviewing the histopathological findings.

## Author Contributions Statement

AKT supervised the study. SG (I) and AKT designed the experiments. SG (I) conducted all the experiments. PS and SC acquired the mass spectrometry data. SG (I) analysed and interpreted the data with critical input from AKT. SSG mentored PS and SG (I) to analyse and interpret the metabolomics data. SG (II) helped to screen the clinical data and acquire biological samples of the study participants. CK validated the histopathological findings. SG (I) and AKT wrote the manuscript. All the authors read, reviewed and approved the final version of the manuscript.

---

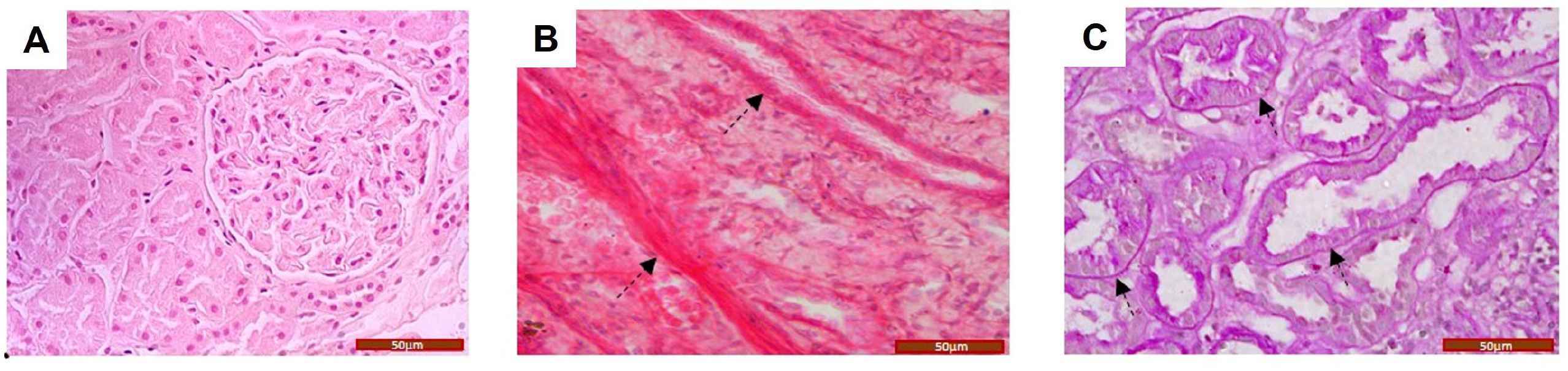

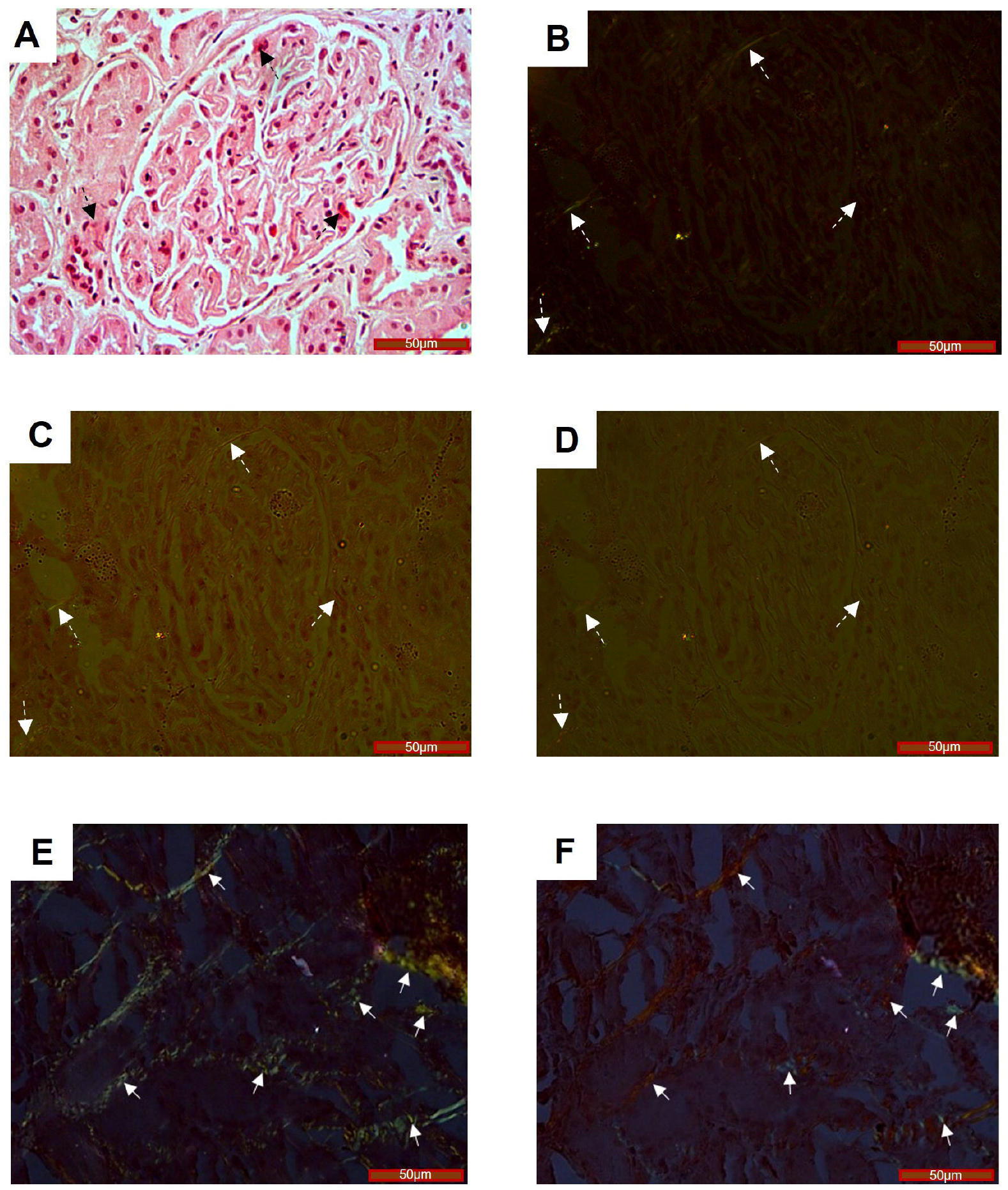

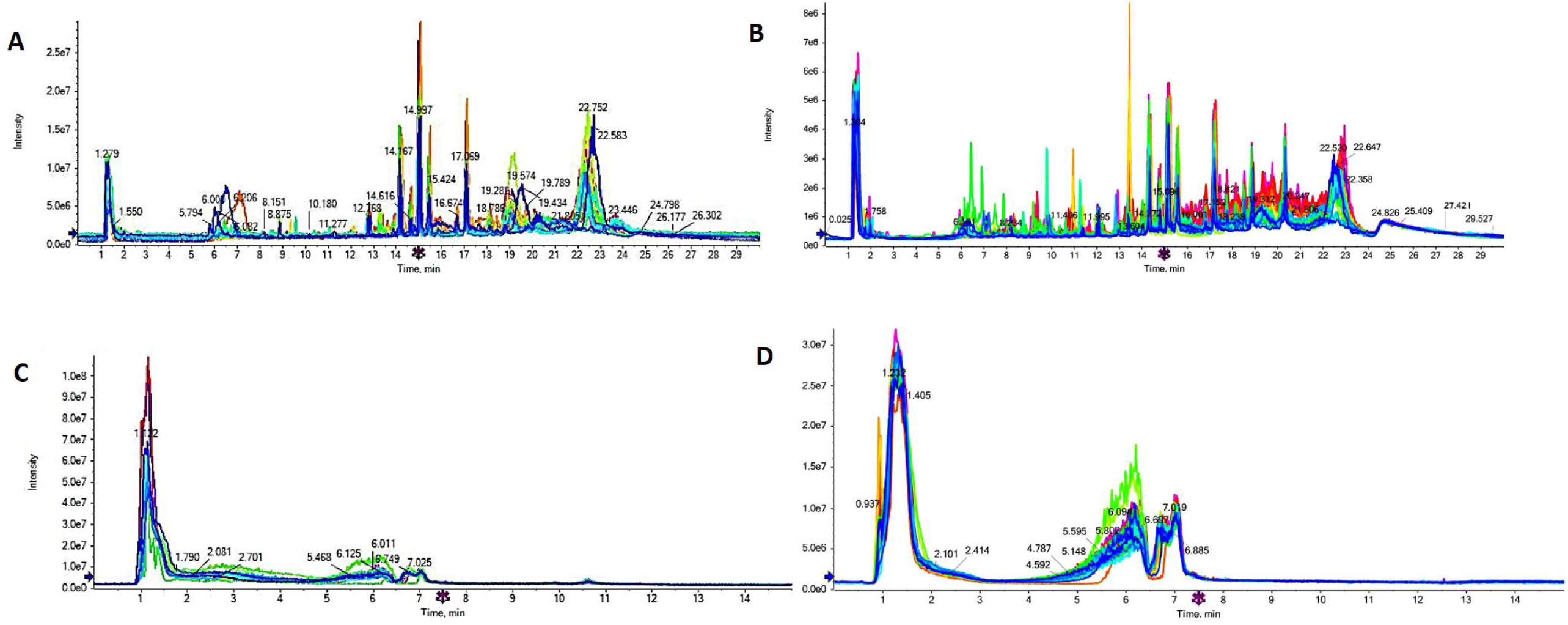

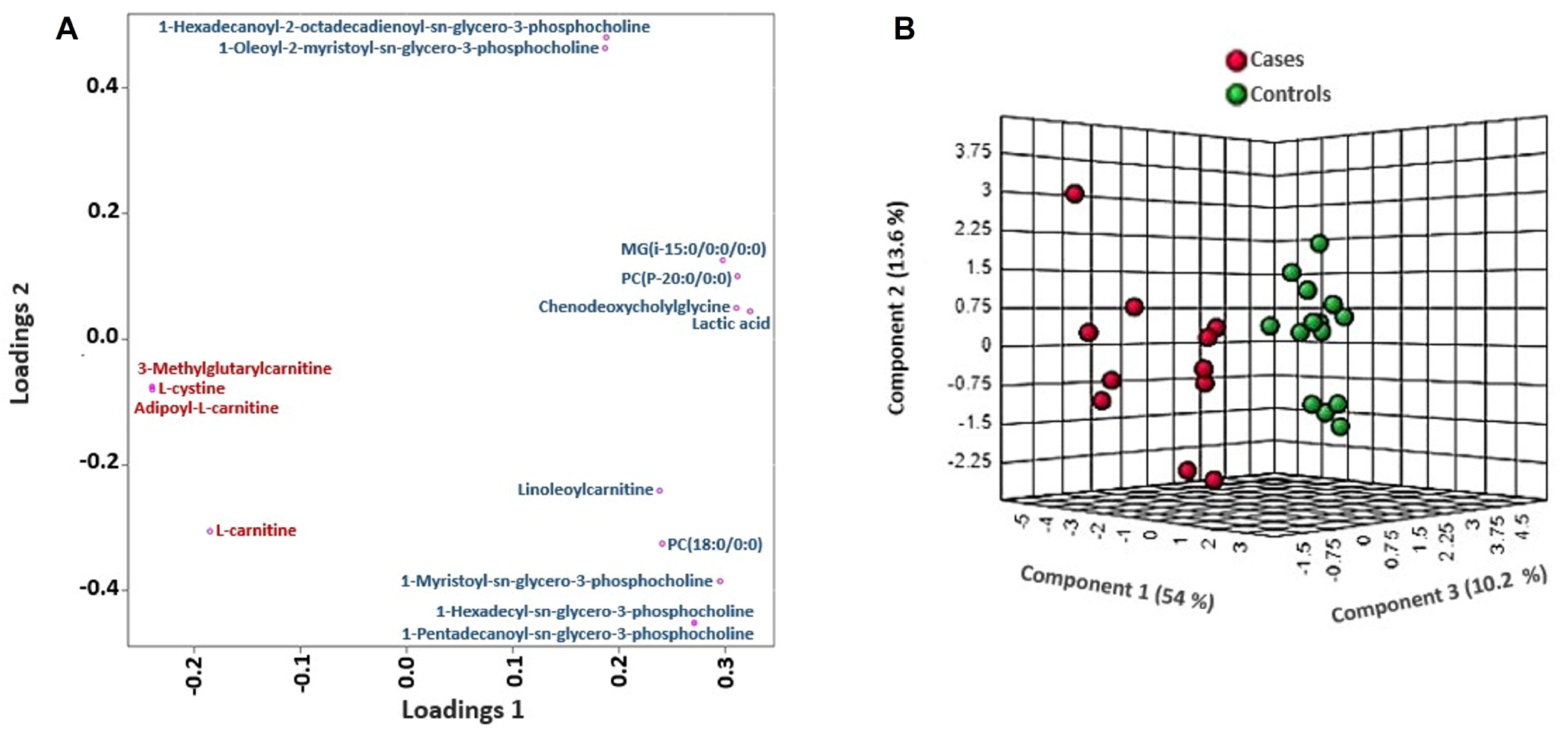

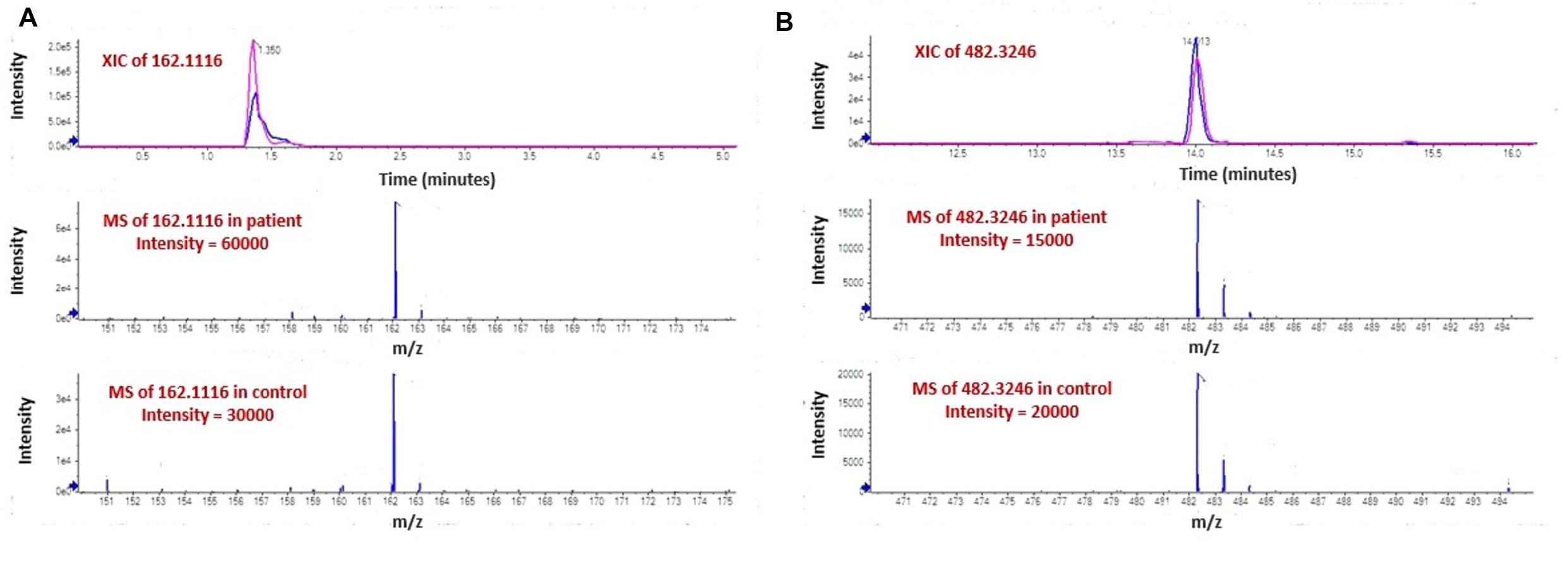

