## Supplementary Information for "Metabolomic fingerprinting for biomarker discovery in renal amyloidosis"

**List of figures**

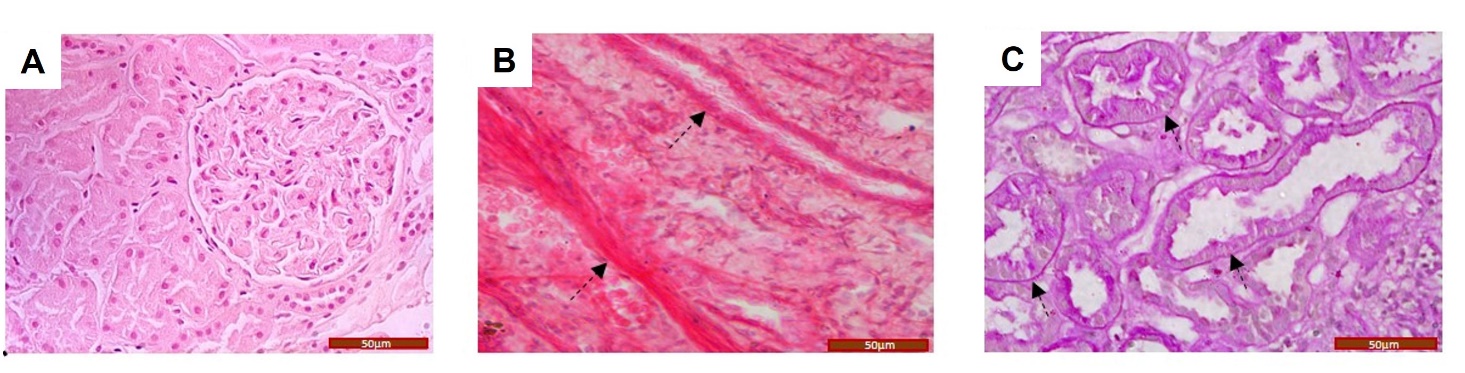
**Figure S1** Histological representation of tissue specimen from a glomerular diseased patient suspected for renal amyloidosis. Diffuse mesangial hypercellularity was observed in the glomerulus (A). Interstitial fibrosis (B) (indicated by black dashed arrows) was evident in the interstitium. Further, distended tubules with thickened basement membranes were seen on the biopsies of these patients (C) (indicated by black dashed arrows). Figure A-B represents H&E-stained image and Figure C represents PAS-stained image. **Scale bar** 50 µm.

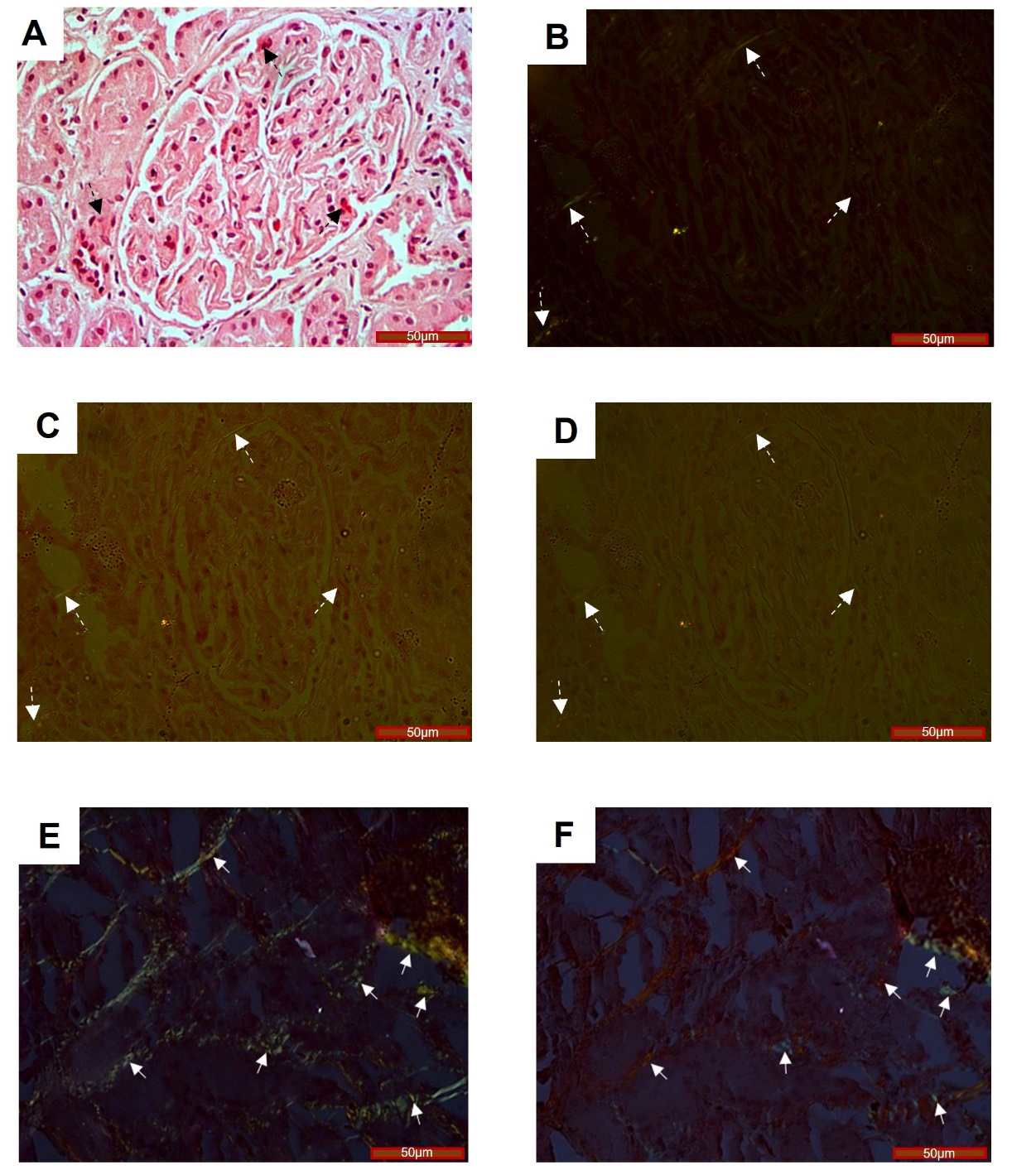

**Figure S2** Representative microscopic images of the Congo red stained renal tissue specimen from a glomerular diseased patient suspected of renal amyloidosis. Hyaline-rich deposits in the glomerulus (A) (indicated by black dashed arrows) exhibited an apple green birefringence (B) (indicated by white dashed arrows) under polarised light. This was further confirmed by the transformation in birefringence colour from apple green to bluish green or reddish-orange upon rotating the polariser by 10º either in a clockwise or anticlockwise direction respectively (C, D) (indicated by white dashed arrows). Additionally, amyloid deposits were also observed in the interstitial spaces (E, F) (indicated by white solid arrows). **Scale bar** 50 µm.

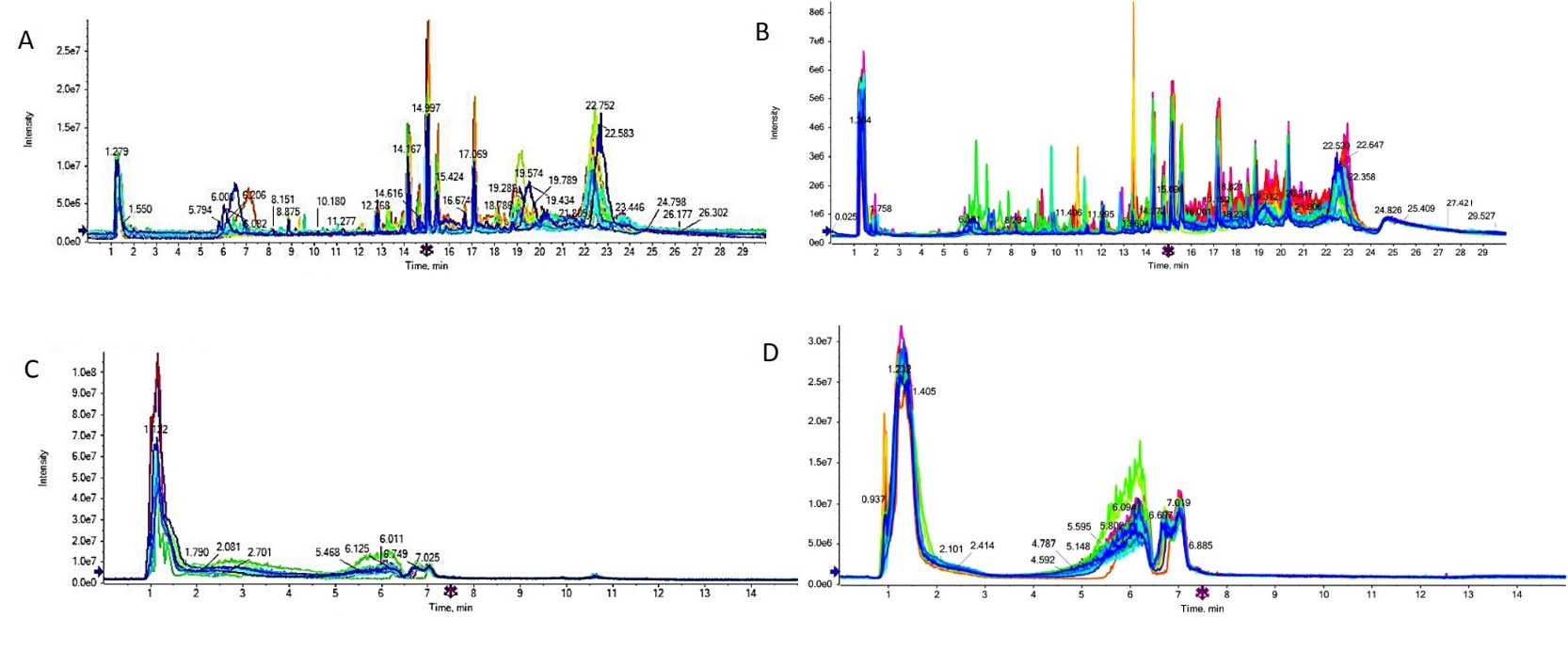

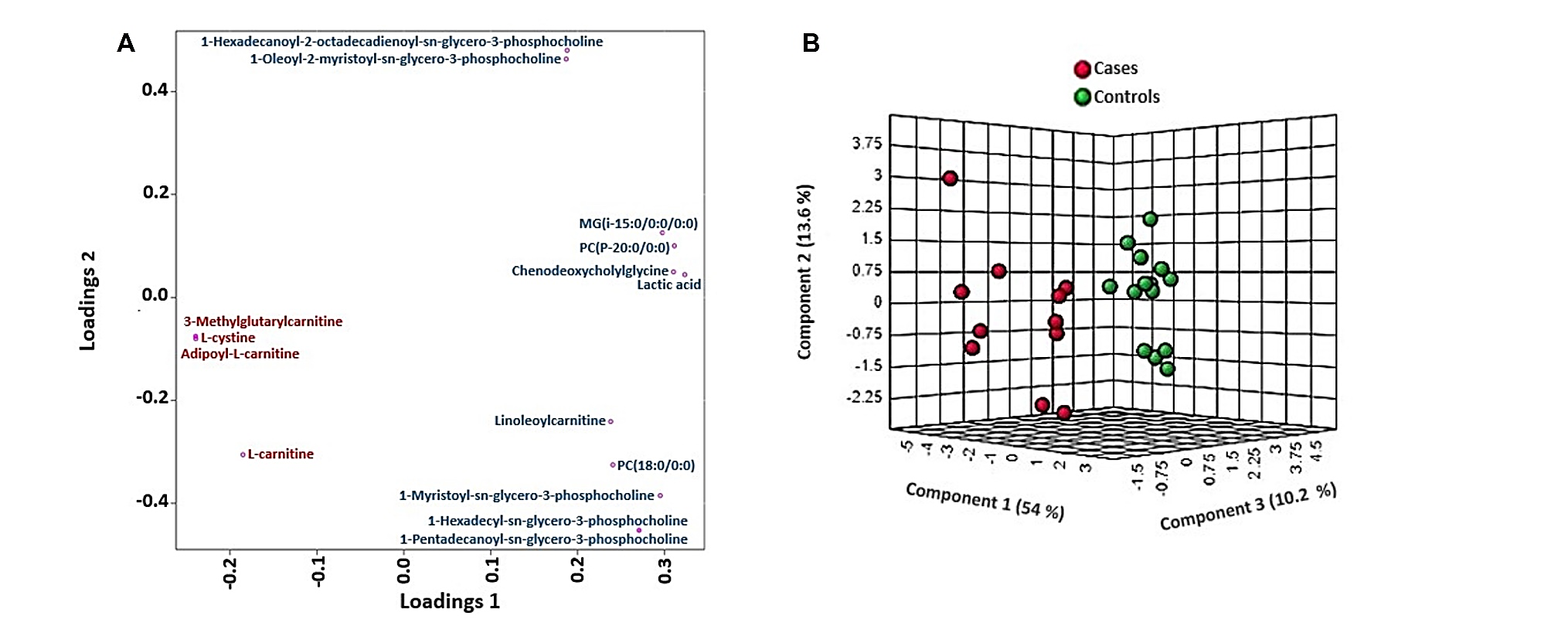
**Figure S3** Representation of overlaid total ion chromatogram in both positive and negative mode of reverse phase (A, B) and HILIC (C, D) LC-MS separation respectively. Each coloured trace represents total chromatogram of each sample respectively.

**Figure S4** PLS-DA loading plot (A) and synchronised 3D plot (B) validating differential distribution of the identified metabolites between the patients and controls. In (A) red colour indicates distribution of upregulated metabolites and blue colour indicates down regulated
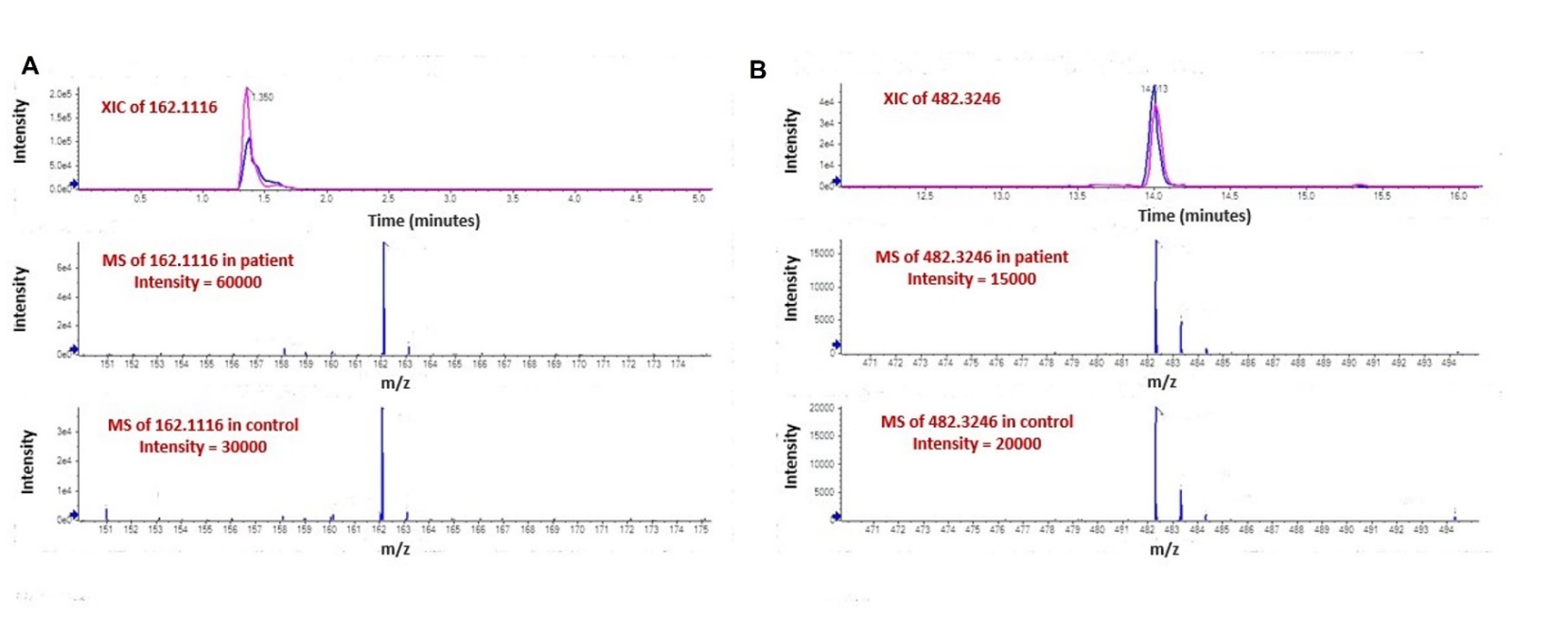
metabolites between cases and controls.

**Figure S5** A representative XIC and MS intensity diagram of an upregulated metabolite, L-carnitine (162.1116) (A) and a down regulated metabolite, 1-Hexadecyl-sn-glycero-3-phosphocholine (482.3246) (B) validating our observations.
